# Protocol for a prospective, multicentre, cross-sectional cohort study to assess personal light exposure

**DOI:** 10.1101/2024.02.11.24302663

**Authors:** Carolina Guidolin, Sam Aerts, Gabriel Kwaku Agbeshie, Kwadwo Owusu Akuffo, Sema Nur Aydin, David Baeza Moyano, John Bolte, Kai Broszio, Guadalupe Cantarero-García, Altug Didikoglu, Roberto Alonso González-Lezcano, Hongli Joosten-Ma, Sofia Melero-Tur, Maria Nilsson Tengelin, María Concepción Pérez Gutiérrez, Oliver Stefani, Ingemar Svensson, Ljiljana Udovicic, Johannes Zauner, Manuel Spitschan

**Affiliations:** Max Planck Institute for Biological Cybernetics, Max Planck Research Group Translational Sensory & Circadian Neuroscience; TUM School of Medicine and Health, Department Health and Sports Sciences, Chronobiology & Health, Technical University of Munich; Research Group Smart Sensor Systems, The Hague University of Applied Sciences; Department of Optometry and Visual Science, Kwame Nkrumah University of Science and Technology; Department of Neuroscience, Izmir Institute of Technology; Research Group ARIE, Department of Architecture and Design, Universidad San Pablo-CEU, CEU Universities; National Institute for Public Health and the Environment of the Netherlands (RIVM); Federal Institute for Occupational Safety and Health (BAuA); Department of Measurement Science and Technology, RISE Research Institutes of Sweden; Lucerne University of Applied Sciences and Arts; TUM Institute for Advanced Study (TUM-IAS), Technical University of Munich

## Abstract

Light profoundly impacts many aspects of human physiology and behaviour, including the synchronization of the circadian clock, the production of melatonin, and cognition. These effects of light, termed the non-visual effects of light, have been primarily investigated in laboratory settings, where light intensity, spectrum and timing can be carefully controlled to draw associations with physiological outcomes of interest. Recently, the increasing availability of wearable light loggers has opened the possibility of studying personal light exposure in free-living conditions where people engage in activities of daily living, yielding findings associating aspects of light exposure and health outcomes, supporting the importance of adequate light exposure at appropriate times for human health. However, comprehensive protocols capturing *environmental* (e.g., geographical location, season, climate, photoperiod) and *individual* factors (e.g., culture, personal habits, behaviour, commute type, profession) contributing to the measured light exposure are currently lacking. Here, we present a protocol that combines smartphone-based experience sampling (experience sampling implementing Karolinska Sleepiness Scale, KSS ratings) and high-quality light exposure data collection at three body sites (near-corneal plane between the two eyes mounted on spectacle, neck-worn pendant/badge, and wrist-worn watch-like design) to capture daily factors related to individuals’ light exposure. We will implement the protocol in an international multi-centre study to investigate the environmental and socio-cultural factors influencing light exposure patterns in Germany, Ghana, Netherlands, Spain, Sweden, and Turkey (minimum n=15, target n=30 per site, minimum n=90, target n=180 across all sites). With the resulting dataset, lifestyle and context-specific factors that contribute to healthy light exposure will be identified. This information is essential in designing effective public health interventions.

## Introduction

### Background

Light exposure significantly affects human health and behaviour, modulating sleep, cognitive functions and neuroendocrine processes [1]. The effects of light on human physiology can be observed acutely as changes in subjective alertness, mood, sleep architecture, heart rate and suppression of the hormone melatonin, usually produced by the brain before the biological night [2], [3], [4], [5], [6]. Furthermore, environmental light acts as the main “zeitgeber” (German for “time giver”) for our circadian system, synchronising endogenous physiological processes to the environmental 24-hour light-dark cycle so that bodily functions are temporally organised to match environmental stimuli [1].

Human circadian rhythms have developed under the stable, naturally occurring 24-hour light-dark cycle, characterised by bright days and dark nights. However, in modern, industrialised societies, we often lack the “dark nights” and the “bright days” that our circadian rhythms adapted for, as individuals can effectively turn night into day with the use of electric light [7]. The advent of light emitting diode (LED) lamps, favourable for their affordable price and energy saving properties, together with the availability of emissive displays such as smartphones and laptops, has played an important role in increasing accessibility and use of electric light [8]. Furthermore, modern lifestyles are characterised by considerable amount of time spent indoors, with estimates reporting up to 90% of indoor time, thus implying insufficient exposure to daylight [9]. These aspects of the modern lifestyle and lighting landscape have caught particular attention from researchers, who are actively investigating the health consequences of aberrant light exposures.

Epidemiological studies have drawn associations between evening light exposure and breast cancer risk, leading to evening light together with insufficient daylight being classified as a human carcinogen [10]. Similarly, cross-sectional and prospective longitudinal studies have shown that presence of light in the sleep environment is linked to higher obesity risk, a known risk factor for cardiovascular disease [11], [12]. New evidence is also demonstrating the role of wrongly timed light exposure for the development of mood disorders [13].

Altogether, the current evidence highlights the need to promote healthy light exposure in public health agendas. Recently, Golombek and colleagues [14] have proposed the notion of “sleep capital”, defined as the compound social, economic and health gain derived, among other factors, by healthy light exposure, consisting of bright (day)light exposure during the day and lack of light at night. As the authors argue, investing in sleep capital by adopting interventions which include adjusting light exposure is necessary for a healthy and productive society and could have profound economic implications, such as increased productivity, cognitive performance, reduced accident rate, and better overall brain health [14]. In terms of defining the appropriate levels of light exposure, recent efforts have been undertaken by an interdisciplinary expert group. Drawing together evidence from laboratory and field studies, Brown and colleagues [15] have provided a framework for understanding the appropriate light amounts for healthy, day-active individuals during daytime, evening, and nighttime hours to maintain optimal physiology and circadian health.

### Measuring determinants of light exposure in the real-world

Given the existing recommendations for optimal light exposure, an important question to address is whether the real-world light exposure patterns experienced by an individual, known as their “spectral diet” [16] actually meet these recommendations, and if this is not the case, what are the daily behaviours and contexts hindering appropriate light exposure? Light exposure in free-living conditions can be measured using wearable devices, known as light loggers, that are worn in various positions on the body, including the wrist (e.g. as a wristwatch), chest (e.g. as a pendant or brooch), or eye level (e.g. on a pair of glasses) by study participants [17]. When worn continuously over time, these wearable devices approximate the retinal irradiance an individual receives daily. The melanopic retinal irradiance drives the physiological effects of light [18]. Light exposure patterns can yield light metrics, including time spent above a specific light threshold (time above threshold, TAT; [19]) and variability of light timing (mean light timing, MLiT; [20]), which can subsequently be linked to health outcomes of interest and compared to the recommended light levels [15], [18].

As wearable light loggers become more accessible, research on light exposure patterns in free-living conditions has surged [17]. Most of the literature, however, remains descriptive, linking light metrics to one or two health outcomes of interest, or showing that individuals indeed largely fail to meet the recommended light levels [21]. While highly informative, these investigations fail to capture the contextual and behavioural dimensions leading to a given light exposure pattern. As proposed by Biller and colleagues [22], an individual’s light exposure profile ultimately depends on many factors, including environmental (geographical location, sunshine hours, climate, temperature and photoperiod), cultural (customs, festivities and norms) and behavioural (lifestyle choices such as commute type and profession, as well as individual preferences) ones, which together interact with the built environment (different window and glazing types, lighting design and architecture).

Importantly, while some determinants of daily light exposure are independent of the individual (e.g. type of lights present in one’s office, type of windows and glazing), individuals can exert a level of control on their light exposure by actively seeking or avoiding behaviours which involve specific light exposure (e.g. having lunch break outside or inside) [23], [24]. Considering the growing evidence that well-timed light exposure is crucial to support human health, it is vital not only to describe the timing and quantities of light that individuals receive during the day but also to understand which contextual and behavioural factors contribute to specific light exposure patterns. Once these have been identified, target behaviours and barriers that prevent optimal light exposure can be addressed by delivering precision behavioural health interventions in simple and accessible ways, such as using mobile apps and chatbots [22].

Here, we outline a comprehensive study protocol for field studies to collect rich and high-quality datasets comprising of light exposure data and its contextual and behavioural contributors. To obtain clean light exposure data from the light loggers, we describe in detail how to instruct participants and ensure their compliance with the protocol. Additionally, we present a questionnaire structure designed to capture daily factors linked to individual light exposure using a mobile app interface. Overall, this protocol provides a framework that researchers interested in collecting light exposure data can flexibly adjust. We will use this protocol to create a reference dataset that characterises individual light exposure over seven days at six different geographical locations in Europe and Africa. Our dataset will characterise light exposure and probe the suitability of light logging devices in different geographical and sociocultural contexts. This will help identify context- and lifestyle-specific factors associated with healthy light exposure patterns, which will serve as a first step to designing effective public health interventions.

## Objectives

The three objectives of the study are

1. To characterise individuals’ light exposure over seven days utilizing a near-corneal-plane light logger placed at the centre of non-prescription glasses frame, along with a light logger as a chest-worn pendant and a wrist-worn light logger;
2. To collect data across six countries (Germany, Ghana, Netherlands, Spain, Sweden, and Turkey); and
3. To investigate the correlations between personal light exposure, physiological variables such as chronotype and light sensitivity, and behavioural outcomes including exercise, mood, and alertness.

## Methods and materials

### Sample

#### Geographical sites and research centres involved

The novelty of this study is that personal light exposure data will be collected across various geographical locations using the same, harmonised protocol. We aim to leverage the collaboration between the following research centres to collect data in six countries: Federal Institute for Occupational Safety and Health (BAuA) and Technical University of Munich (TUM) in Germany, Kwame Nkrumah University of Science and Technology (KNUST) in Ghana, The Hague University of Applied Sciences (THUAS) in the Netherlands, Fundación Universitaria San Pablo CEU (FUSP-CEU) in Spain, Research Institutes of Sweden (RISE) in Sweden, and the Izmir Institute of Technology (IZTECH) in Turkey. We believe that the diversity in culture, latitude, photoperiod, climate, built environment and, hence, light exposure behaviours between these six locations, will provide interesting insights for the objectives of this study.

#### Participant recruitment

Participants will be recruited by self-selection through advertisements which will be posted at the local hubs as well as in local newsletters. Participants interested in the study will be directed to an online platform (Research Electronic Data Capture; REDCap) [25], [26] for the initial screening survey. Detailed information about the study and its aim will be provided during this screening step. Inclusion and exclusion criteria (**Table 1**) will be tested using a questionnaire on the same online platform. This questionnaire will also collect demographic data (age, sex, gender, native language(s) and occupational status). If eligible for the study, participants will then be contacted by the experimenters to agree on possible participation dates and discuss any further questions.

**Table 1:** Inclusion and exclusion criteria.

| Aspect | Assessment modality | Exclusion criterion and cut-off | Timing of Screening |
| --- | --- | --- | --- |
| Age | Self-report | <18 years<br>>65 years | Initial screening survey |
| Psychiatric and sleep disorders | Self-report | Presence of any | Initial screening survey |
| Tobacco and recreational drug use | Self-report | Regular use (1/week or more) | Initial screening survey |
| Medication intake | Self-report | Presence of any known to influence photosensitivity | Initial screening survey |
| Visual acuity | Self-report | Requirement of prescription glasses during the experimental week | Initial screening survey |
| Normal vision | Self-report | History of ocular or retinal diseases, colour blindness | Initial screening survey |
| Location during experimental week | Self-report | Exiting local hubs (≥60 km) during weekdays (Monday to Friday) of the experimental week | Initial screening survey |
| Shift work | Self-report | Shiftwork in the past two months | Initial screening survey |
| Parenthood | Self-report | Parent of a child <1 year old | Initial screening survey |
| Full-time employment | Self-report | Unemployment, leave, working part-time (<80%), studying | Initial screening survey |

Furthermore, they will be sent a picture of what the light logger looks like and asked if they feel comfortable wearing them throughout the experimental week. They will also be informed about the availability of the researchers throughout the experiment in case of doubts or technical issues with the light logger. Participants will be compensated at the end of the study according to their compliance with the experimental procedure: for every day of wearing the light logger for at least 80% of their waking hours (as defined by the Munich Chronotype Questionnaire; MCTQ) [27], volunteers will receive financial compensation, such that those adhering to the whole duration of the experiment will receive more than those adhering, for example, to only four out of the seven experimental days. The rates of financial compensation will depend on each measurement site and local customs. Data collection can terminate after reaching at least n=15 per site, with a target of n=30. The researchers will terminate the study for an individual participant in case of technical issues which do not allow the experiment to continue, e.g., when the light logger is not working as expected.

#### Inclusion and exclusion criteria

Eligible participants will be selected according to the inclusion and exclusion criteria listed in **Table 1**. These include demographic as well as mental and physical health parameters. Individuals with corrected vision requiring prescription glasses during the experimental week will be excluded due to incompatibility with our light glasses. However, individuals with a) prescription lenses or b) prescription glasses but are able and willing to wear prescription contact lenses during the experimental week will be able to participate in our study. Individuals suffering from psychiatric or sleep disorders will be excluded from the study. Furthermore, intake of any drugs and/or medications known to influence photosensitivity will be considered a criterion for exclusion. Finally, only people based at or near (<60 km) the local hubs of each geographical location during the weekdays (Monday to Friday) of the experiment will be accepted for this study to have similar environmental conditions across participants at each measurement hub. All criteria mentioned above for inclusion and exclusion will be assessed by self-report through REDCap [25], [26]. The eligibility criteria used here can be modified for studies in which the goal is to assess a different population.

### Protocol

#### Study design

This experiment is an observational field study in which all participants at the six different sites will undergo the same experimental conditions and questionnaires. These are shown in **Table 2**.

**Table 2:** Measurement schedule.

| Read-out | Measurement modality | Sampling frequency | Timing of sampling | N per participant |
| --- | --- | --- | --- | --- |
| **Objective individual light exposure** | Light logger | Continuous measurement over 7 days | Every 10 seconds | Approx. 10080 |
| **Objective activity/rest** | Actimeter | Continuous measurement over 7 days | Every 10 seconds | Approx. 10080 |
| **Chronotype** | Munich Chronotype Questionnaire (MCTQ, circadian time) and Morning Evening Questionnaire (MEQ, circadian preference) | 1 measurement over 7 days | First experimental day | 1 |
| **Subjective sleep** | Consensus Sleep Diary (CSD) | 7 measurements over 7 days | Every morning | 7 |
| **Subjective hourly light exposure and activities** | Modified Harvard Light Exposure Questionnaire (modified H-LEA) | 7 measurements over 7 days | Every evening | 7 |
| **Subjective wellbeing** | WHO-5 wellbeing index (WHO-5) | 7 measurements over 7 days | Every evening | 7 |
| **Exercise frequency and type** | Exercise log | 7 measurements over 7 days | Every evening | 7 |
| **Subjective light exposure** | Modified Harvard Light Exposure Questionnaire (modified H-LEA). Experience sampling: punctual measurement on participants' current light conditions | 24 measurements over 7 days | 4 times/day | 22 |
| **Subjective alertness** | Karolinska Sleepiness Scale (KSS). Experience sampling: punctual measurement on participants' current light conditions | 22 measurements over 7 days | 4 times/day | 22 |
| **Subjective mood** | MoodZoom questionnaire | 22 measurements over 7 days | 4 times/day | 22 |
| **Experience log** | Custom-made questionnaire and open-ended questions about positive and negative experiences wearing the light logger | Continuous measurement over 7 days | Throughout the experiment | Depending on participant |
| **Wear log** | Custom-made questionnaire about time of taking the device off and putting it back on | Continuous measurement over 7 days | Throughout the experiment | Depending on participant |
| **Subjective light sensitivity** | Visual Light Sensitivity Questionnaire 8 (VLSQ-8) | 1 measurement over 7 days | Last experimental day | 1 |
| **User experience of wearing the light logger** | Open-ended questions | 1 measurement over 7 days | Last experimental day | 1 |
| **Sleep environment** | Assessment of sleep environment questionnaire (ASE) | 1 measurement over 7 days | Last experimental day | 1 |
| **Subjective light exposure** | Light Exposure Behaviour Assessment (LEBA) | 1 measurement over 7 days | Last experimental day | 1 |

#### Procedure

A schematic representation of the experimental procedure is illustrated in **Figure 1**. Eligible participants will start the experiment on a Monday with an in-person visit to the office or laboratory of the selected hub and finish the experiment on the following Monday. On the first Monday (day 1), they will receive a detailed explanation of the experiment and sign an informed consent document. Volunteers will then be provided with three wearable light loggers to be worn at the near-corneal plane, at the chest level and at the wrist. They will receive detailed instructions on using both devices correctly, including removing them when in contact with water and during contact sports. Participants will also install the MyCap app [28], which integrates with REDCap and is used to fill in daily questionnaires and set alarms on their phones as reminders to complete the scheduled questionnaires on the app. Before leaving, participants will complete two questionnaires measuring circadian time and circadian preference (Munich Chronotype Questionnaire; MCTQ and Morning Eveningness Questionnaire; MEQ).

**Figure 1:**
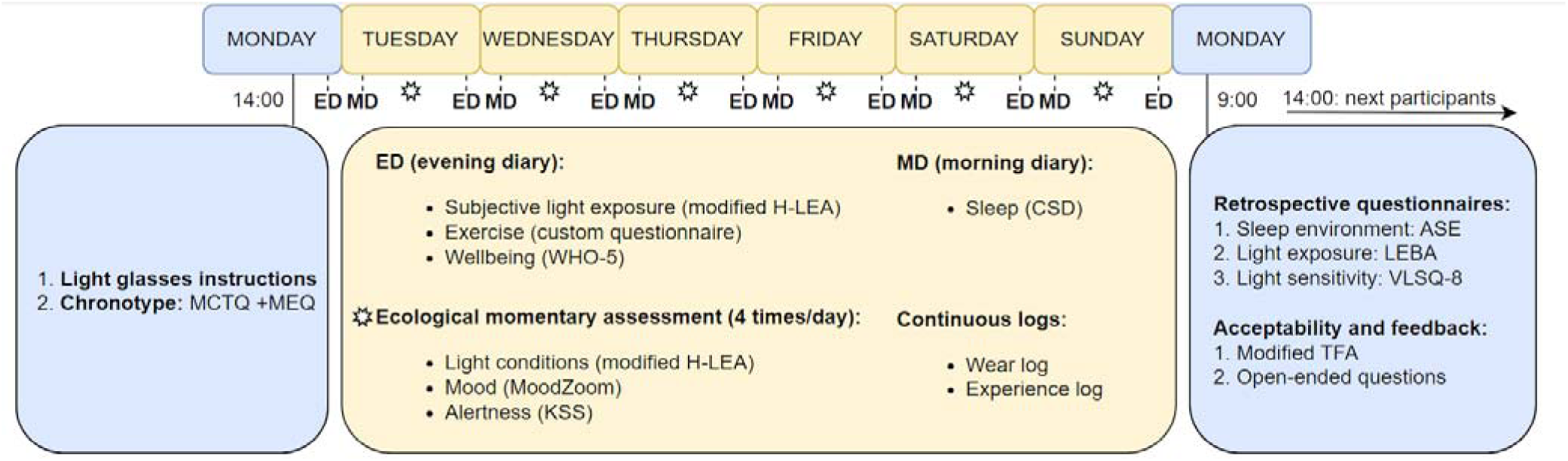
Schematic representation of the experimental timeline of the experiment (Monday to Monday).

Participants will wear the three light loggers throughout the week during their daily activities. Participants will be instructed to log non-wear time as follows. When taking off the spectacle-mounted light logger during the day, they will press the event button on the device and place it in a black bag. They will then log this action in the “Wear log” on the MyCap app. Similarly, when putting the light logger back on, they will take it out of the black bag, press the event button, and log this action in the “Wear log” on the MyCap app. If the participants forget the black bag, they will be prompted to describe where they placed the light glasses instead. Before sleep, participants will place the spectacle-worn light logger facing upwards on a bedside table or flat surface near their bed. They will also log this action in the “Wear log”. In case participants exit the local area (defined as a 60 km radius from the local hub), they will describe where they are located at this time and report when they re-entered the local area on the “Wear log”. In case they forget to log an activity, participants are allowed to log any of the five possible “Wear log” events (“Light logger on”, “Light logger off”, “Light logger off before sleep”, “Exiting local area”, “Re-entering local area”) as “past events” which happened previously (see Appendix).

Every morning after waking up, participants will start wearing the light logger and log this in the “Wear log” on the MyCap app. They will also fill in a questionnaire regarding their sleep (Consensus Sleep Diary; CSD). Throughout the day, they will receive notifications at four scheduled times to fill in questionnaires regarding their current light conditions (modified Harvard Light Exposure Assessment Questionnaire; modified H-LEA), alertness (Karolinska Sleepiness Scale; KSS) and mood (MoodZoom). In the evening, participants will complete questionnaires about their light exposure and activities during the last 24 hours (modified H-LEA), wellbeing (WHO-5 Wellbeing Index; WHO-5) and exercise (custom questionnaire). Throughout the experiment, participants will also report their positive and negative experiences in the “Experience log” (see Appendix).

Participants will return to the local centre on the following Monday, one week after the experiment starts. On this day, they will return the devices and complete a retrospective questionnaire regarding their light exposure (Light Exposure Behaviour Assessment; LEBA), light sensitivity (Visual Light Sensitivity Questionnaire-8; VLSQ-8), and their sleep environment (Assessment of Sleep Environment questionnaire; ASE) during the seven days they participated in the study. Furthermore, they will complete open-ended questions about their opinions on the light logger device (see Appendix). After completing these questionnaires, participants will be reimbursed based on their compliance with the experiment. The devices will be charged, and the next set of participants will start the experiment later that day.

### Measures

In this section, we provide detailed information on the objective and subjective health-related measures collected in this study and described in *Procedure*.

### Objective health-related measures

#### Personal light logging

To measure personal light exposure, we will deploy ActLumus light loggers (Condor Instruments, São Paulo, Brazil) worn by participants for one week. ActLumus light loggers contain ten spectral channels, the outputs of which are combined to estimate photopic and melanopic irradiance. Throughout the trial, participants will wear three light loggers:

1. To measure light centrally in the near-corneal plane, the light loggers will be placed on the frame of non-prescription glasses. A 3D-printed holder for the light loggers has been designed and attached to the bridge of the glasses frame, enabling the insertion and removal of the ActLumus devices.
2. To measure light on the chest, the light loggers will be clipped to clothing or worn as a pendant.
3. To measure light on the wrist, a conventional location, the light loggers will be worn with manufacturer-provided wrist bands.

The choice of having three light loggers instead of only one is a technical one. Currently, there is no “best practice” for which measurement level (eye, chest, or wrist) is most accurate, or whether the three are comparable. For this reason, we collect light exposure data at all three levels, with the intent of exploring how the measured illuminance compares between them. The sampling interval of each ActLumus light logger will be set to 10 seconds to achieve highly temporally resolved data, and the devices will never be turned off nor charged during the experimental week. Light exposure data for each participant will then be downloaded only upon the return of the devices on the final Monday (day 8). The choice of light loggers used here can vary depending on the availability.

As the use of non-prescription glasses still requires the use of lenses without optical power, the transmittance properties of the lenses will be measured between 250 and 2500 nm.

### Activity measurement

One of the ActLumus light loggers will be worn on the wrist. The ActLumus measures movement through an integrated tri-axial accelerometer and is used in field studies such as ours to distinguish wake and sleep time. Participants will be instructed to keep the wrist-worn device on during the day and night and only remove it when in contact with water and during contact sports.

### Physiological, behavioural and contextual determinants of personal light exposure

To understand which physiological, behavioural and contextual factors contribute to participants’ objectively measured light exposure, we will collect a variety of subjectively reported health-related measures that can provide insights into subjects’ daily activities and environments. This information will be collected at study intake, throughout the study or at discharge in the form of questionnaires through the REDCap/MyCap interface, as described in *Procedure* (refer to **Figure 1** for the frequency and timing of each measurement).

### Physiological measures

#### Chronotype questionnaires

On the first day of the experiment, participants will complete two questionnaires measuring circadian time and circadian preference: the Munich Chronotype Questionnaire (MCTQ, [27]) and the Morning-Eveningness Questionnaire (MEQ, [29]). The MCTQ is used to assess circadian time using questions about their sleep and wake habits during work and free days and commute type. The MEQ is used to determine the circadian preference of individuals to perform certain activities at specific times of the day.

#### Visual Light Sensitivity Questionnaire-8 (VLSQ-8)

Participants complete the 8-point Visual Light Sensitivity Questionnaire-8 (VLSQ-8; [30]) at study discharge to answer questions about their visual light sensitivity during the experimental week. The questions include aspects of frequency and severity of photosensitivity as well as impacts of photosensitivity on daily behaviours, and participants answer using a 5-point Likert scale (1 = “Never” to 5 = “Always”).

### Behavioural measures

#### Morning sleep log

Every morning after waking up, participants fill in the core Consensus Sleep Diary [31] consisting of 9 items to assess their sleep timing, sleep duration during the night, and subjective sleep quality. This last item is scored on a five-point Likert scale (1 = “Very poor” to 5 = “Very good”).

#### Ecological momentary assessment (“Current conditions”)

Four times a day (at 11:00, 14:00, 17:00 and 20:00), participants fill in a questionnaire concerning their current light conditions, mood and sleepiness. The researcher sends a reminder message through the REDCap/MyCap messaging channel, and phone alarms set by participants at these times serve to ensure compliance. Firstly, current light conditions are tested through a multiple-choice question, where participants can choose one of 8 possible light scenarios as the “main light source” and, if applicable, as the “secondary light source”. The potential light sources to choose from consist of the same categories listed in the modified Harvard Light Exposure Assessment diary, which participants fill in every evening (see “Light exposure and activity diary”, H-LEA; [32]). Secondly, a modified MoodZoom questionnaire [33] assesses current mood. Lastly, sleepiness is assessed using the Karolinska Sleepiness Scale (KSS; [34]) on a 10-point Likert scale ranging from 1 = “Extremely alert” to 10 = “Extremely sleepy, fighting sleep”.

#### Exercise log

Every evening before sleep, participants complete a custom-made questionnaire about the exercise they performed during the day. This questionnaire was designed to assess intensity (vigorous/moderate/light, lack of exercise) and location (indoors/outdoors) of exercise, as well as sedentary time (“How much time did you spend sitting or reclining?”).

#### Wellbeing log

Every evening before sleep, participants complete a modified version of the WHO-5 Wellbeing Index [35], consisting of 5 statements (1 = “I have felt cheerful and in good spirits”, 2 = “I have felt calm and relaxed”, 3 = “I have felt active and vigorous”, 4 = “How would you rate the quality of your sleep last night?”, and 5 = “My daily life has been filled with things that interest me”). Participants have to express agreement using a 5-point Likert scale ranging from 0 = “At no time” to 5 = “All of the time” (for statements 1, 2, 3 and 5) and from 1 = “Very poor” to 5 = “Very good” for statement 4.

#### Worktime log

Every evening before sleep, participants complete a custom-made questionnaire on the clock times they went to their workplace, how, and when they returned home.

#### Light exposure and activity log

Every evening, participants have to fill in a modified version of the Harvard Light Exposure Assessment (H-LEA; [32]). This is referred to as “mH-LEA” and is done on paper using a form provided by the experimenter during the in-person visit (see Appendix). Participants are asked to report, for each hour of the day, the primary light source they are exposed to and the activity they performed in that hour. The primary light source is described as “the biggest and brightest light source”. They can choose between 8 light categories (”Electric light source indoors (e.g., lamps such as LEDs)”, “Electric light source outdoors (e.g., street lights)”, “Daylight indoors (through windows)”, “Daylight outdoors (including being in the shade)”, “Emissive displays (e.g., smartphone, laptop etc.)”, “Darkness (outdoors and/or indoors)”, “Light entering from outside during sleep (e.g., daylight, street lights etc.)”). If they believe they are exposed to a combination of lights within the same hour, they can choose from a list of possible combinations. With regards to their activity, they could choose between 8 categories (“Sleeping in bed”, “Awake at home”, “On the road with public transport/car”, “On the road with bike/on foot”, “Working in the office/from home”, “Working outdoors (including lunch break outdoors), “Free time outdoors (e.g. garden/park etc.), “Other: please specify (e.g. sport)”. To ensure that participants complete this task, they send a picture of the completed form every night and upload it to a shared folder (separate for each participant) where the experimenter could check compliance. Furthermore, they are asked to rate the confidence in their answers on MyCap, where they can answer using a 5-point Likert scale ranging from 1 = “Not confident at all” to 5 = “Completely confident”.

#### Light Exposure Behaviour Assessment (LEBA)

The 22-item Light Exposure Behaviour Assessment (LEBA; [23]) is used to retrospectively assess individuals’ light behaviours during the experimental week at study discharge. Since the first three items of this instrument ask questions related to wearing blue-filtering, orange-tinted and/or red-tinted glasses, which do not apply to our participants due to the presence of the light logger device, these items are eliminated. The final questionnaire thus comprises the remaining 19 items. These concern specific behaviours such as exposure to daylight, smartphone use, light-related bedtime habits and electric light use at home. Participants can express the frequency of such behaviours using a 5-point Likert scale ranging from 1 = “Never” to 5 = “Always”.

### Contextual measures

#### Assessment of Sleep Environment (ASE) questionnaire

The 13-item Assessment of Sleep Environment (ASE) questionnaire is used to ask participants about aspects such as light, noise, temperature and humidity in their sleeping environment [36], which might affect their sleeping quality as well as the light measured by the light logger placed next to participants during sleep (e.g., in case of light coming through windows during sleep). Participants can express their agreement to each item using a 5-point Likert scale (1 = “Strongly agree” to 5 = “Strongly disagree”).

#### Environmental light logging

To measure the environmental light in the local site during each experimental week, one ActLumus light logger will optionally be placed on the rooftop of a chosen building. The set-up for these environmental light measurements consists of a black metal floor, where the device lies horizontally, covered by a plastic half-dome to minimise light scattering while ensuring protection from the elements. This set-up is placed on the rooftop before participants start the study every week and remains there for the entire week until participants discharge, measuring environmental light with a sampling interval of 30 seconds. Each day, a researcher will check and, if necessary, clean the outside and/or the inside of the set-up from dirt or rain. At the end of each experimental week, the data from this environmental light logger will be downloaded, and the device will be charged before being placed back on the rooftop just before the next participants start the study on the same day. When such measurements are impossible, secondary data sources, including historical weather data, sunshine duration, sunrise/sunset times, or existing radiation measurement infrastructure, will be used.

#### Translation and adaptation of questionnaires

To run the study in our five sites, translation of surveys and questionnaires is required. To this end, a team-based, multi-step process will be employed to achieve this goal, involving a diverse group of individuals, including trained translators and experts in the survey’s subject matter (based on the “TRAPD” approach to translate questionnaires). The source language is English and the target languages are German (Germany), Dutch (Netherlands), Swedish (Sweden), Spanish (Spain) and Turkish (Turkey). In Ghana, the original English version is used. A detailed description of the strategy used to translate the questionnaires in reported in the Appendix.

#### Trial feasibility

The current protocol was trialled in an independent data collection effort taking place from August to November 2023 in Tübingen, Germany. A total of 26 participants (14 female; mean age±1SD: 28.0±5.2) worn a corneal-plane light logger (ActLumus) and a wrist-worn actigraphy and light logger (ActTrust2) for a week (Monday to Monday), and completed the same subjective health-related measures described in the current protocol. The protocol was found to be largely feasible, and feedback from the participants was taken on in refining the protocol presented here. Furthermore, successful strategies for ensuring data quality throughout the experiment as well as during data analysis were documented and will be implemented in the current protocol. This will ensure a standardised data curation and analysis approach across the six geographical locations. The data collected in this independent data collection campaign will be published independently of this protocol.

## Statistical analysis

### Power analysis

A sample size calculation based on power analysis was performed based on a framework described elsewhere [37]. The calculation was based on a historical dataset [38] provided by one of the geographical locations (Germany: BAuA); where participants measured light exposure for multiple days in winter, spring, and summer with devices attached to clothing at chest height. A suitable subset of this data was used to calculate the necessary sample size to reach a power of 0.8 across common light exposure metrics when comparing them between winter and summer seasons. While the experiment producing the historical data deviates somewhat from the current study’s experimental structure, it still allows for a realistic comparison of metrics between different environmental conditions while considering intra-individual variability. The sample size calculation is based on a bootstrap resampling of daily metrics between winter and summer. For each resampled dataset, significance was tested in a mixed-effect model (fixed effect: season, random effect: participants) with a significance level of 0.05. The fraction of significant differences was compared against the power level threshold of 0.8. The required sample size is the minimum sample size that reaches this threshold, with 1000 resamples per sample size (sample sizes from 3 to 50 were tested). A total of twelve metrics were analyzed:

- Geometric mean of melanopic EDI (lx)
- Geometric standard deviation of melanopic EDI (lx)
- Luminous exposure (lx\*h)
- Time above 250 lux (h)
- Time above 1000 lux (h)
- Mean timing of light above 250 lux (h)
- Mean timing of light below 10 lux (h)
- Intradaily variability
- Mean across the darkest (L5) hours (lx)
- Midpoint of the darkest (L5) hours (lx)
- Mean across brightest (M10) hours (lx)
- Midpoint of the brightest (M10) hours (lx)

Three metrics had no effect in the historical dataset and thus did not reach the power threshold (Geometric standard deviation, mean timing of light above 250 lux, midpoint of darkest 5 hours). With a sample size of 15 participants, eight out of nine metrics showed sufficient power (intradaily variability: 21 participants to threshold power). Even considering a high dropout rate of 33% leaves seven out of nine metrics sufficiently powered (mean of darkest 5 hours: 15 participants to threshold power).

### Pre-processing

Objectively measured light exposure data will be log-transformed (base 10) to account for large light level differences, such as indoor and outdoor light exposure.

Data from the light logger will be processed to separate non-wear time from wear time. For this purpose, the Wear log will be considered the “ground truth” in terms of detection of non-wear time. Once confirmed by visual inspection, non-wear times of ≥5 minutes will be removed.

We will apply stringent exclusion criteria for our confirmatory tests (see Confirmatory analysis). We will exclude the following missing data in hourly analyses:

- Missing entry on the modified H-LEA for a given hour during waking hours: if no category has been selected for a given waking hour (waking hours as specified in the sleep log of the corresponding day);
- Non-wear times of 50% for a given hour.

Furthermore, we will exclude an individual day from the analysis if 20% of the objective light exposure data from a participant’s waking hours (specified in the MCTQ) is missing. This does not apply to the first and last experimental days, as these are not “complete” days (participants will receive and return the light logger throughout the day).

When data have been excluded from confirmatory analyses, we may include them in future exploratory analyses.

### Statistical analysis and pre-processing

We plan to analyse all data with the R software and the package LightLogR (https://tscnlab.github.io/LightLogR/index.html) which provides a workflow for the processing, visualization and metrics calculation based on wearable light logger data. If not otherwise specified below, the planned method for statistical analysis is through (linear) mixed-effect models implemented with the lme4 package [39]. Equations follow the notation used by the package. p-values obtained by likelihood-ratio tests of the full model with the effect in question, against the model without the effect. p-values less than or equal to 0.05 will be considered significant. p-value adjustment for multiple tests within each hypothesis is planned using Benjamini and Hochberg’s *false discovery rate* (FDR) method [40].

### Confirmatory analysis

We plan to perform the three following confirmatory analyses:

1. H1: We hypothesize that there is a significant relationship between hourly self-reported light exposure categories and hourly median objective light exposure.

a. Preparation: Hourly entry on light sources from daily modified H-LEA will serve as categorical variables. In the case of two light sources for a given hour, only the primary light source will be considered (as reported by participants). The median melanopic equivalent daylight illuminance (melanopic equivalent daylight illuminance; mEDI) as measured objectively by the light logger for the corresponding hour will be calculated.
b. Analysis: mel EDI is used as the dependent variable, and H-LEA as the fixed effect, participants within each geolocation as random effect. Participant’s geolocation, sex, age, occupational status and chronotype (MCTQ) are added as covariates. The dependency of mel EDI and H-LEA as well as the weekday is allowed to vary between participants within a geolocation. The resulting formula is as follows: 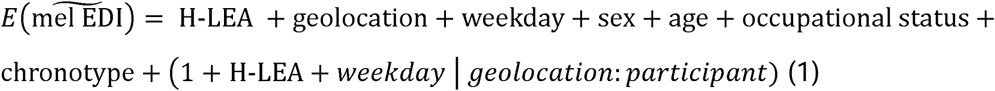
2. H2: We hypothesize that MCTQ-measured chronotype MSF_sc_ (mid-sleep on free days corrected for sleep debt on weekdays) and MLiT^250^ ^lx^ ^mEDI^ (mean light timing >250 lx melanopic EDI) are correlated, such that earlier chronotypes receive light earlier in the day.

a. Preparation: calculate MCTQ-derived MSF_sc_ for each participant and calculate MLiT^250^ ^lx^ ^mEDI^ as average clock time of all data points >250 lx mEDI over the six measurement days for each participant.
b. Analysis: Spearman’s rank correlation coefficient between MSF_sc_ and 6-day average MLiT^250^ ^lx^ ^mEDI^ for each participant. Additional models with various ring-fenced covariates will be built in future steps.
3. H3: We hypothesize that there is a significant difference between daily average objective light exposure and geographical location, and additionally, that the differences in photoperiod explain a substantial part of that relationship

a. Preparation: calculate the daily mel EDI light dose (in lx*h) as measured objectively by the light logger for the corresponding day. Calculate also the photoperiod of that day for a given location as the time from sunrise until sunset (sun elevation equal to zero), as calculated by the sun angles given from the *oce* R package [41].
b. Analysis: daily mel EDI light dose (in lx*h)is the dependent continuous variable. Geolocation is the independent categorical variable. A second step also includes photoperiod. Weekday, sex, age, and chronotype are covariates. Participant ID within geolocation is a random effect, as is the weekday effect for each participant. The full formula is as follows: 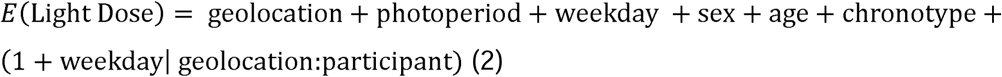

### Exploratory analyses

We plan to explore several relationships regarding physiological, behavioural, and contextual self-reported data. These are listed here for transparency.

1. Light exposure

1.1. Relationship between metrics of light exposure that describe light properties (melanopic EDI and photopic illuminance) and between metrics describing the temporal pattern of light exposure, including light regularity index (LRI), intraday variability (IV), interday stability (IS).
1.2. Comparison between objectively measured personal light exposure during weekdays and weekends.
1.3. Relationship between retrospective light exposure items as measured by the LEBA instrument and objective light exposure.
1.4. Relationship between environmental conditions during the experimental week (e.g. photoperiod availability, sunlight hours and temperature) and objective personal light exposure.
1.5. Relationship between objective personal light exposure measured and mood and alertness ratings measured throughout the day.
1.6. Relationship between subjective light sensitivity as reported by the VLSQ-8 and objective personal light exposure.
1.7. Relationship between daily objective personal light exposure and wellbeing scores as measured by the WHO-5 questionnaire.
1.8. Relationship between exercise frequency and type as measured by the exercise log and objective personal light exposure.
1.9. Relationship between geolocation, photoperiod and other metrics of light exposure (see 1.1), also in interaction with the weekday.
2. Light logger acceptability:

2.1. Descriptive analysis of open-ended questions on wearing the light logger.
2.2. Relationship between negative and positive experiences as reported in the experience log and non-wear time as reported in the wear log.

## Data storage and privacy

Data collected through REDCap and MyCap will be pseudonymized in the system and stored on this system until the end of data analysis. Anonymised data will be made publicly available after the publication of the primary research publication.

## Outcome measures

### Primary outcome measures

Our primary outcome measures are listed in **Table 3**. They include objectively measured daily light exposure (examined in H1 and H3) and chronotype (examined in H2).

**Table 3:** Primary outcome measures.

| Measurement modality | Derived measure and unit | Definition | Number of measurements per participant | Pre-processing | Linked confirmatory analysis |
| --- | --- | --- | --- | --- | --- |
| Objective light exposure at three sites | Melanopic EDI (lux) | Weighted spectral irradiance | Depending on participant | 1. Removal of non-wear times $\geq 10$ minutes after visual inspection and if probability score between non-wear sources $\geq 0.66$<br>2. Removal of single day if 20% data is missing during given day between Tuesday and Sunday<br>3. Removal of participant if missing data for entire day between Tuesday and Sunday | H1, H2 & H3 |
| Subjective light exposure | Rating – different modified H-LEA categories | Perceived light exposure | 7 | 1. Removal of hours where entry is missing<br>2. Exclusion of secondary light source for hourly each entry | H1 |
| Chronotype | MCTQ | Chronotype | 1 | Calculation on $MSF_{sc}$ | H2 |

### Secondary outcome measures

Our secondary outcome measures will be described using summary statistics and explored in exploratory analyses. Specifically, we will explore the relationship between objectively measured personal light exposure, physiological variables (chronotype and light sensitivity), behavioural variables (such as exercise, mood, and alertness) and contextual variables (sleep environment).

## Code, data and materials availability

Upon conclusion of the primary analyses, the data will be made available under the Creative Commons license (CC-BY) with no reservations in the supplementary material of the research publication and/or on a public repository (e.g., FigShare).

## Additional files

GuidolinEtAl_2024_Appendix.pdf: Supplementary document containing additional information on data collection, and translation/adaptation of questionnaires used in the study.

## Ethics approval

This research protocol was reviewed and approved by the Medical Ethics Committee of the Technical University of Munich (2023-115-S-KK).

## Conflicts of interest

The authors declare that the study will be conducted without any financial or commercial relationships that could be explained as a conflict of interest.

## Author contributions

*Conceptualisation*: C.G., M.S.

*Data curation*: n/a

*Formal Analysis*: n/a

*Funding acquisition*: L.U., D.B.M., M.N.T., O.S., M.S.

*Investigation*: n/a

*Methodology*: C.G., L.U., K.B., D.B.M., S.M.-T., G.C-G., R.A.G.-L., S.A., J.B., H.J.-M., M.N.T., O.S., A.D., J.Z., M.S.

*Project administration*: M.S.

*Resources*: n/a

*Software*: C.G., J.Z., M.S.

*Supervision*: n/a

*Validation*: n/a

*Visualisation*: C.G.

*Writing – original* draft: C.G., M.S.

*Writing – review & editing*: C.G., S.A., G.K.A., K.O.A., S.N.A., R.A.G.-L., D.B.M., J.B., K.B., G.C.-G., A.D., H.J.-M., S.M.-T., M.N.T., M.C.P.G., O.S., I.S., L.U., J.Z., M.S.

## Funding statement

The project (22NRM05 MeLiDos) has received funding from the European Partnership on Metrology, co-financed by the European Union’s Horizon Europe Research and Innovation Programme and by the Participating States. Further finding is provided by the TUM Global Incentive Fund (GIF0000031) to support the collaboration between TUM and KNUST, and TÜBİTAK –The Scientific and Technological Research Council of Türkiye (Grant no. 224S740).

## Notes

### Competing Interest Statement

The authors have declared no competing interest.

### Summary of Updates

Major revisions of the manuscript.

## References

[1] C. Blume, C. Garbazza, and M. Spitschan, “Effects of light on human circadian rhythms, sleep and mood,” Somnologie, vol. 23, no. 3, pp. 147–156, Sep. 2019, doi: 10.1007/s11818-019-00215-x.

[2] Cibeira, et al., “Healthcare | Free Full-Text | Bright Light Therapy in Older Adults with Moderate to Very Severe Dementia: Immediate Effects on Behavior, Mood, and Physiological Parameters.” Accessed: Aug. 02, 2023. [Online]. Available: https://www.mdpi.com/2227-9032/9/8/1065

[3] R. Lok, K. C. H. J. Smolders, D. G. M. Beersma, and Y. A. W. de Kort, “Light, alertness, and alerting effects of white light: a literature overview,” J Biol Rhythms, vol. 33, no. 6, pp. 589–601, Dec. 2018, doi: 10.1177/0748730418796443.

[4] S. L. Chellappa, M. C. M. Gordijn, and C. Cajochen, “Can light make us bright? Effects of light on cognition and sleep,” in Progress in Brain Research, vol. 190, Elsevier, 2011, pp. 119–133. doi: 10.1016/B978-0-444-53817-8.00007-4.

[5] S. L. Chellappa, R. Lasauskaite, and C. Cajochen, “In a Heartbeat: Light and Cardiovascular Physiology,” Frontiers in Neurology, vol. 8, 2017, Accessed: Dec. 04, 2023. [Online]. Available: https://www.frontiersin.org/articles/10.3389/fneur.2017.00541

[6] C. B. Swope et al., “Factors associated with variability in the melatonin suppression response to light: A narrative review,” Chronobiology International, pp. 1–15, Mar. 2023, doi: 10.1080/07420528.2023.2188091.

[7] S. L. Chellappa, “Individual differences in light sensitivity affect sleep and circadian rhythms,” Sleep, vol. 44, no. 2, p. zsaa214, Feb. 2021, doi: 10.1093/sleep/zsaa214.

[8] S. Westland, Q. Pan, and S. Lee, “A review of the effects of colour and light on non-image function in humans,” Coloration Technology, vol. 133, no. 5, pp. 349–361, 2017, doi: 10.1111/cote.12289.

[9] N. E. Klepeis et al., “The National Human Activity Pattern Survey (NHAPS): a resource for assessing exposure to environmental pollutants,” J Expo Sci Environ Epidemiol, vol. 11, no. 3, Art. no. 3, Jul. 2001, doi: 10.1038/sj.jea.7500165.

[10] K. Y. Lai, C. Sarkar, M. Y. Ni, L. W. T. Cheung, J. Gallacher, and C. Webster, “Exposure to light at night (LAN) and risk of breast cancer: A systematic review and meta-analysis,” Science of The Total Environment, vol. 762, p. 143159, Mar. 2021, doi: 10.1016/j.scitotenv.2020.143159.

[11] Y. Cho, S.-H. Ryu, B. R. Lee, K. H. Kim, E. Lee, and J. Choi, “Effects of artificial light at night on human health: A literature review of observational and experimental studies applied to exposure assessment,” Chronobiology International, vol. 32, no. 9, pp. 1294–1310, Oct. 2015, doi: 10.3109/07420528.2015.1073158.

[12] Y.-M. M. Park, A. J. White, C. L. Jackson, C. R. Weinberg, and D. P. Sandler, “Association of Exposure to Artificial Light at Night While Sleeping With Risk of Obesity in Women,” JAMA Internal Medicine, vol. 179, no. 8, pp. 1061–1071, Aug. 2019, doi: 10.1001/jamainternmed.2019.0571.

[13] A. C. Burns et al., “Day and night light exposure are associated with psychiatric disorders: an objective light study in >85,000 people,” Nat. Mental Health, vol. 1, no. 11, pp. 853–862, Nov. 2023, doi: 10.1038/s44220-023-00135-8.

[14] D. A. Golombek et al., “Sleep Capital: Linking Brain Health to Wellbeing and Economic Productivity Across the Lifespan,” The American Journal of Geriatric Psychiatry, Jul. 2024, doi: 10.1016/j.jagp.2024.07.011.

[15] T. M. Brown et al., “Recommendations for daytime, evening, and nighttime indoor light exposure to best support physiology, sleep, and wakefulness in healthy adults,” PLoS Biol, vol. 20, no. 3, p. e3001571, Mar. 2022, doi: 10.1371/journal.pbio.3001571.

[16] F. S. Webler, M. Spitschan, R. G. Foster, M. Andersen, and S. N. Peirson, “What is the ‘spectral diet’ of humans?,” Current Opinion in Behavioral Sciences, vol. 30, pp. 80–86, Dec. 2019, doi: 10.1016/j.cobeha.2019.06.006.

[17] S. Hartmeyer, F. Webler, and M. Andersen, “Towards a framework for light-dosimetry studies: Methodological considerations,” Lighting Research & Technology, p. 14771535221103258, Jul. 2022, doi: 10.1177/14771535221103258.

[18] M. Spitschan et al., “Verification, analytical validation and clinical validation (V3) of wearable dosimeters and light loggers,” Digit Health, vol. 8, p. 20552076221144858, 2022, doi: 10.1177/20552076221144858.

[19] S. Hartmeyer and M. Andersen, “Towards a framework for light-dosimetry studies: Quantification metrics,” Lighting Research & Technology, p. 147715352311705, May 2023, doi: 10.1177/14771535231170500.

[20] K. J. Reid, G. Santostasi, K. G. Baron, J. Wilson, J. Kang, and P. C. Zee, “Timing and intensity of light correlate with body weight in adults,” PLoS One, vol. 9, no. 4, p. e92251, 2014, doi: 10.1371/journal.pone.0092251.

[21] A. Didikoglu et al., “Associations between light exposure and sleep timing and sleepiness while awake in a sample of UK adults in everyday life,” Proceedings of the National Academy of Sciences, vol. 120, no. 42, p. e2301608120, Oct. 2023, doi: 10.1073/pnas.2301608120.

[22] A. M. Biller, P. Balakrishnan, and M. Spitschan, “Behavioural determinants of physiologically-relevant light exposure,” Mar. 18, 2024, *OSF*. doi: 10.31219/osf.io/xpt4e.

[23] M. A. Siraji et al., “An inventory of human light exposure behaviour,” Sci Rep, vol. 13, no. 1, p. 22151, Dec. 2023, doi: 10.1038/s41598-023-48241-y.

[24] M. A. Siraji, M. Spitschan, V. Kalavally, and S. Haque, “Light exposure behaviors predict mood, memory and sleep quality,” Sci Rep, vol. 13, no. 1, p. 12425, Aug. 2023, doi: 10.1038/s41598-023-39636-y.

[25] P. A. Harris, R. Taylor, R. Thielke, J. Payne, N. Gonzalez, and J. G. Conde, “Research electronic data capture (REDCap)—A metadata-driven methodology and workflow process for providing translational research informatics support,” Journal of Biomedical Informatics, vol. 42, no. 2, pp. 377–381, Apr. 2009, doi: 10.1016/j.jbi.2008.08.010.

[26] P. A. Harris et al., “The REDCap consortium: Building an international community of software platform partners,” Journal of Biomedical Informatics, vol. 95, p. 103208, Jul. 2019, doi: 10.1016/j.jbi.2019.103208.

[27] T. Roenneberg, L. K. Keller, D. Fischer, J. L. Matera, C. Vetter, and E. C. Winnebeck, “Human activity and rest in situ,” Methods Enzymol, vol. 552, pp. 257–283, 2015, doi: 10.1016/bs.mie.2014.11.028.

[28] P. A. Harris et al., “MyCap: a flexible and configurable platform for mobilizing the participant voice,” JAMIA Open, vol. 5, no. 2, p. ooac047, Jul. 2022, doi: 10.1093/jamiaopen/ooac047.

[29] J. A. Horne and O. Östberg, “Individual differences in human circadian rhythms,” Biological Psychology, vol. 5, no. 3, pp. 179–190, Sep. 1977, doi: 10.1016/0301-0511(77)90001-1.

[30] J. D. Verriotto et al., “New Methods for Quantification of Visual Photosensitivity Threshold and Symptoms,” Transl Vis Sci Technol, vol. 6, no. 4, p. 18, Jul. 2017, doi: 10.1167/tvst.6.4.18.

[31] C. E. Carney et al., “The consensus sleep diary: standardizing prospective sleep self-monitoring,” Sleep, vol. 35, no. 2, pp. 287–302, Feb. 2012, doi: 10.5665/sleep.1642.

[32] A. Bajaj, B. Rosner, S. W. Lockley, and E. S. Schernhammer, “Validation of a light questionnaire with real-life photopic illuminance measurements: the Harvard Light Exposure Assessment questionnaire,” Cancer Epidemiol Biomarkers Prev, vol. 20, no. 7, pp. 1341–1349, Jul. 2011, doi: 10.1158/1055-9965.EPI-11-0204.

[33] A. Tsanas et al., “Daily longitudinal self-monitoring of mood variability in bipolar disorder and borderline personality disorder,” J Affect Disord, vol. 205, pp. 225–233, Nov. 2016, doi: 10.1016/j.jad.2016.06.065.

[34] T. Akerstedt and M. Gillberg, “Subjective and objective sleepiness in the active individual,” Int J Neurosci, vol. 52, no. 1–2, pp. 29–37, May 1990, doi: 10.3109/00207459008994241.

[35] Bech, P., “Measuring the dimensions of psychological general well-being by the WHO-5.,” QoL Newsletter, vol. 32, pp. 15–16, 2004.

[36] M. A. Grandner et al., “Development and Initial Validation of the Assessment of Sleep Environment (ASE): Describing and Quantifying the Impact of Subjective Environmental Factors on Sleep,” Int J Environ Res Public Health, vol. 19, no. 20, p. 13599, Oct. 2022, doi: 10.3390/ijerph192013599.

[37] J. Zauner, L. Udovicic, and M. Spitschan, “Power analysis for personal light exposure measurements and interventions,” In Review, preprint, Dec. 2023. doi: 10.21203/rs.3.rs-3771881/v1.

[38] L. L. A. Price, M. Khazova, and L. Udovičić, “Assessment of the Light Exposures of Shift-working Nurses in London and Dortmund in Relation to Recommendations for Sleep and Circadian Health,” Annals of Work Exposures and Health, vol. 66, no. 4, pp. 447–458, Apr. 2022, doi: 10.1093/annweh/wxab092.

[39] D. Bates, M. Mächler, B. Bolker, and S. Walker, “Fitting Linear Mixed-Effects Models Using **lme4**,” J. Stat. Soft., vol. 67, no. 1, 2015, doi: 10.18637/jss.v067.i01.

[40] Y. Benjamini and Y. Hochberg, “Controlling the False Discovery Rate: A Practical and Powerful Approach to Multiple Testing,” Journal of the Royal Statistical Society: Series B (Methodological), vol. 57, no. 1, pp. 289–300, Jan. 1995, doi: 10.1111/j.2517-6161.1995.tb02031.x.

[41] D. Kelley and C. Richards, oce: Analysis of Oceanographic Data. 2024. [Online]. Available: https://dankelley.github.io/oce/

