## Appendix for "Protocol for a prospective, multicentre, cross-sectional cohort study to assess personal light exposure"

Carolina Guidolin[1, 2]
Sam Aerts[3]
Gabriel Kwaku Agbeshie[4]
Kwadwo Owusu Akuffo[4]
Sema Nur Aydin[5]
Roberto Alonso González-Lezcano[6]
David Baeza Moyano[6]
John Bolte[3, 7]
Kai Broszio[8]
Guadalupe Cantarero-García[6]
Altug Didikoglu[5]
Hongli Joosten-Ma[3]
Sofia Melero-Tur[6]
Maria Nilsson Tengelin[9]
María Concepción Pérez Gutiérrez[6]
Oliver Stefani[10]
Ingemar Svensson[9]
Ljiljana Udovicic[8]
Johannes Zauner[2]
Manuel Spitschan[1, 2, 11 *]

1 Max Planck Institute for Biological Cybernetics, Max Planck Research Group Translational Sensory & Circadian Neuroscience

2 TUM School of Medicine and Health, Department Health and Sports Sciences, Chronobiology & Health, Technical University of Munich

3 Research Group Smart Sensor Systems, The Hague University of Applied Sciences

4 Department of Optometry and Visual Science, Kwame Nkrumah University of Science and Technology

5 Department of Neuroscience, Izmir Institute of Technology

6 Research Group ARIE, Department of Architecture and Design, Universidad San Pablo-CEU, CEU Universities

7 National Institute for Public Health and the Environment of the Netherlands (RIVM)

8 Federal Institute for Occupational Safety and Health (BAuA)

9 Department of Measurement Science and Technology, RISE Research Institutes of Sweden

10 Lucerne University of Applied Sciences and Arts

11 TUM Institute for Advanced Study (TUM-IAS), Technical University of Munich, Garching, Germany

### Additional measures collected

This section includes details on additional subjective measures collected during the experiment which do not relate to the physiological, behavioural and contextual determinants of light exposure.

#### Wear log

Throughout the day, participants are instructed to report their wear and non-wear time (only concerning the spectacle-worn glasses) in a digital log book. Specifically, they have five choices of wear log entry: 1 = “Taking the light glasses off”, 2 = “Putting the light glasses on”, 3 = “Taking the light glasses off before sleep and placing them on a nightstand or flat surface”, 4 = “Leaving <study location > and its surroundings (60 km radius)” and 5 = “Re-entering < study location > and its surroundings (60 km radius)”. For options 1 to 3, participants also press the button on the light glasses to signal an event occurring, and in the case of 1, they are asked to confirm whether they place the light glasses in the black bag provided to them and if they are in movement. Options 4 and 5 are introduced to control for potential differences in personal light exposure due to environmental availability rather than behaviour. For all the five wear log entry choices, participants must state whether they are logging a present or a past event.

Additionally, participants are asked for the reasons they took the light logger off (“What is prompting you to remove the light logger?”), with the options “Sports activity”, “Leisure activity where I do not feel comfortable wearing the light logger (e.g. public space)”, “Activity involving contact with water (e.g., showering or bathing)”, “Discomfort due to wearing the light glasses (e.g. disturbance to eyesight or pain due to weight)” and “Other (please specify)”.

#### Experience log

During the week, participants also have the opportunity to report their experiences with the light glasses using a log on the app. This log prompts participants to describe situations in which they received verbal or nonverbal feedback from others and personal comfort with wearing the light glasses. They are also asked whether and how these experiences might influence their future use of the light glasses.

#### Open-ended questions

To further probe the usability of our light logger, we present participants with the following open-ended questions: “Can you describe any challenges or discomfort you experienced while wearing the light glasses? How did you cope with them?”, “In what situations did you notice the light glasses having the most impact on your daily activities or behaviour?”, “How did you adapt your behaviour, if at all, because of the light glasses? Please provide some examples.”, “Can you share any suggestions or improvements for the design or functionality of the light glasses (comprising the sensor and the glasses) for future experiments?”, “Please describe any situations or activities where the light glasses failed to capture your "typical" light exposure because you had to take them off?”, and “How comfortable were the light glasses for you to wear during your daily activities?”.

#### Translation and adaptation of questionnaires

The following section describes in detail the strategy used to translate the questionnaires in the different target languages, based on the “TRAPD” approach:

**Team approach:**

- Assemble a translation team of individuals from diverse backgrounds, bringing together interdisciplinary expertise.
- Ensure that the team consists of at least three independent members (two acting as translators and one as a reviewer/adjudicator).

**Translator selection:**

- Ideally, choose two translators with experience (and some training) in translating surveys/questionnaires.
- The translators should have proficiency in both the source language (the language of the original questionnaire) and the target language (the language into which the questionnaire will be translated). Ideally the translators translate into their mother tongue.
- Ideally at least one of the two translators is a trained and/or professional translator.

**First project meeting**:

- Discuss potential future challenges translating the questionnaires and surveys and flag items that may be difficult (source questionnaire).

**Initial parallel translations:**

- Begin the translation process with two parallel translations of the source instrument into the target language (by the two translators mentioned above). The two translators should not contact each other while translating, but delivers independent translations.

**Review discussion and adjudication**

- In a “Review” discussion, all items of the questionnaire should be discussed at least by the three persons of the translation team, possibly more:  compare and discuss the two initial translations and try to agree on one translation: this may be one of the two initial translations, a blend of both or a completely new translation, developed during the discussion.
- A “Reviewer” should chair the session. This should be an expert in the matter with good proficiency in both English and the target language.
- Include an adjudicator with expert knowledge in the subject matter to resolve any discrepancies or ambiguities in the translations. This may be the third person in the translation team, the

"Reviewer" (then called "Reviewer-cum-Adjudicator"), or an additional, 4th person. If the adjudicator is a 4th person, he/she may participate in the Review meeting, or be consulted after the meeting.

- The translators should be present during this session to answer any language-related questions, clarify, and bring in the translation perspective.

**Pre-test of the translation:**

- Conduct a cognitive pretesting (n≈10) of the translated questionnaire to assess its comprehensibility and cultural appropriateness.
- This pretest should involve a sample of the target population who will eventually complete the translated questionnaire.
- Based on the pretest results, consider making necessary adjustments to the translated questionnaire to improve its clarity and cultural relevance.
- If significant issues arise, conducting further cognitive pretesting iterations is advisable.
- In case the cognitive pretests reveal weaknesses of the source questionnaire, please report back to the central team.

**Final Review:**

- Review and finalise the translated questionnaire based on the feedback and insights gained from the run-through and cognitive pretesting.

**Quality Assurance:**

- Maintain a comprehensive documentation of the entire translation process, including all versions of the questionnaire, meeting notes, and participant feedback.
- Ensure that the final translated questionnaire is linguistically accurate, culturally appropriate, and equivalent in meaning to the source instrument

Example of the light exposure and activity diary (modified H-LEA) completed by participants

#### LIGHT EXPOSURE AND ACTIVITY DIARY

*Please complete every day before going to bed and upload on the shared folder emailed to you on the first day of the experiment.*

For each hour of the day:

1. Select the light source you were exposed to by choosing from the categories below (first row)
2. Select the activity you were doing by choosing from the categories below (second row)

LIGHT EXPOSURE CATEGORIES:

L: Electric light source indoors (e.g.: lamps such LEDs etc.)

I: Daylight indoors (through windows)

E: Emissive displays (e.g.: smartphone, laptop etc.) S: Electric light source outdoors (e.g.: street lights) O: Daylight outdoors (including being in the shade) D: Darkness during sleep

W: Light entering from outside during sleep (e.g.: daylight, street lights etc.)

If you are exposed to a combination of lights, please choose one from:

L+I, L+E, I+E, S+O, D+W

**ACTIVITY CATEGORIES:**

1: Sleeping in bed

**2. Awake at home**

**3. On the road with public transport/car**

**4. On the road with bike/on foot**

**5. Working in the office/from home**

**6. Working outdoors (including lunch break outdoors)**

**7. Free time outdoors (e.g. garden/park etc.)**

**8. Other: please specify (e.g. sport indoors, sport outdoors)**

**Tip:** start from the first hour when you woke up until the hour when you go to bed.

| | 0:00 - 1:00 | 1:00 - 2:00 | 2:00 - 3:00 | 3:00 - 4:00 | 4:00 - 5:00 | 5:00 - 6:00 | 6:00 - 7:00 | 7:00 - 8:00 | 8:00 - 9:00 | 9:00 - 10:00 | 10:00 - 11:00 | 11:00 - 12:00 | 12:00 - 13:00 |
|---|---|---|---|---|---|---|---|---|---|---|---|---|---|
| Light source | D | D | D | D | D | D | D+W | D+W | I | I+E | I+E | I+E | I+E |
| Activity | 1 | 1 | 1 | 1 | 1 | 1 | 1 | 1 | 2 | 4 | 4 | 4 | 4 |

| | 13:00 - 14:00 | 14:00 - 15:00 | 15:00 - 16:00 | 16:00 - 17:00 | 17:00 - 18:00 | 18:00 - 19:00 | 19:00 - 20:00 | 20:00 - 21:00 | 21:00 - 22:00 | 22:00 - 23:00 | 23:00 - 24:00 |
|---|---|---|---|---|---|---|---|---|---|---|---|
| Light source | O | I | I | I | I | O | O | S | L | L+E | D |
| Activity | 6 | 4 | 4 | 4 | 4 | 7 | 7 | 4 | 2 | 2 | 1 |
